# Reduction in severity of all-cause gastroenteritis requiring hospitalisation in children vaccinated against rotavirus in Malawi

**DOI:** 10.1101/2021.06.21.21259250

**Authors:** Jonathan J. Mandolo, Marc Y. R. Henrion, Chimwemwe Mhango, End Chinyama, Richard Wachepa, Oscar Kanjerwa, Chikondi Malamba-Banda, Isaac T. Shawa, Daniel Hungerford, Arox W. Kamng’ona, Miren Iturriza-Gomara, Nigel A. Cunliffe, Khuzwayo C. Jere

## Abstract

**Background:** Rotavirus is the major cause of severe gastroenteritis in children aged <5 years. Introduction of Rotarix® rotavirus vaccine (RV1) in Malawi in 2012 has reduced rotavirus-associated hospitalisations and diarrhoeal mortality. However, RV1 impact on gastroenteritis severity remains unknown. We conducted a hospital-based surveillance study to assess RV1 impact on gastroenteritis severity in children aged <5 years, in Malawi.

**Methods:** Stool samples were collected from children hospitalised with acute gastroenteritis from December 2011 – October 2019. Gastroenteritis severity was determined using Ruuska and Vesikari scores. Rotavirus was detected in stool using Enzyme Immunoassay. Rotavirus genotypes were determined using nested RT-PCR. Associations between RV1 vaccination and gastroenteritis severity were investigated using adjusted linear regression.

**Results:** In total, 3,159 children were recruited. After adjusting for Mid-Upper Arm Circumference, age, weight, gender and receipt of other vaccines, all-cause gastroenteritis severity scores were 2.21 units lower (95% CI 1.85, 2.56; *p*<0.001) among RV1-vaccinated (*n*=2,224) compared to RV1-unvaccinated children (*n*=935); the decrease was comparable between rotavirus-positive and rotavirus-negative cases in all age groups. The reduction in severity score was observed against every rotavirus genotype, although the magnitude was smaller among those infected with G12P[6] compared to the remaining genotypes (*p*=0.011). Other than RV1 vaccination, age was the only variable associated with gastroenteritis severity. Each one-year increment in age was associated with a decrease of 0.43 severity score (95% CI 0.26, 0.60; *p*<0.001).

**Conclusion:** Our findings provide additional evidence of RV1 impact in a high disease burden, low-income country, lending further support to Malawi’s rotavirus vaccine programme.

**Summary:** In a long-term hospital-based surveillance study in Malawi, we found evidence of the reduction in gastroenteritis severity among hospitalised RV1-vaccinated children infected with both homotypic and heterotypic rotavirus strains and off-target RV1 vaccine effects against non-rotavirus diarrhoeal severity.

## 1. Introduction

Rotavirus is a leading cause of acute gastroenteritis among children worldwide. Despite the introduction of rotavirus vaccines in many countries, rotavirus is still associated with an estimated 128,500 deaths annually. Over 90% of these cases occur in low and middle-income countries (LMICs) in sub-Saharan Africa and South-east Asia [1].

Rotavirus has a segmented double-stranded ribonucleic acid (dsRNA) genome, surrounded by a triple layered capsid. Most human infections are associated with group A rotaviruses [2], which can further be classified using a dual classification system into G and P types, according to their neutralizing antibody response or nucleotide differences of the genes encoding their outer glycoprotein VP7 and protease-sensitive VP4, respectively [3]. At least 36 G types and 51 P types have been reported (https://rega.kuleuven.be/cev/viralmetagenomics/virus-classification). Of these, genotypes G1P[8], G2P[4], G3P[8], G4P[8], G9P[8] and G12P[8] are the most frequent cause of rotavirus disease in humans worldwide [4, 5].

Malawi introduced the G1P[] Rotarix® rotavirus vaccine (RV1) into its national Expanded Programme on Immunization (EPI) schedule on 28^th^ October 2012, with doses administered at 6 and 10 weeks of age, which was followed by a decline in rotavirus-associated hospitalisations [6] and a reduction in gastroenteritis-related mortality [7]. However, the impact of RV1 on the severity of gastroenteritis has not yet been assessed. We conducted an analysis of the severity of gastroenteritis by comparing the Ruuska and Vesikari disease severity scores [8] in children presenting with rotavirus and non-rotavirus laboratory confirmed gastroenteritis before and after RV1 introduction, and between vaccinated and non-vaccinated children post RV1 introduction.

## 2. Materials and methods

### 2.2 Study population

Children under the age of five years who presented with acute gastroenteritis (defined as the passage of at least three looser-than-normal stools in a 24-hour period for less than seven days duration) were enrolled at the Queen Elizabeth Central hospital (QECH), Blantyre, which is the main referral hospital for the southern region of Malawi. To assess the impact of RV1 introduction on the severity of gastroenteritis, data were examined from children hospitalised with gastroenteritis before (December 2011 to October 2012) and after (November 2012 to October 2019) RV1 introduction. These populations were used to (i) assess the impact of RV1 on the severity of all-cause gastroenteritis; and (ii) determine whether gastroenteritis severity differed by rotavirus genotype in RV1-vaccinated and RV1-unvaccinated children.

### 2.3 Clinical and demographic variables

Gastroenteritis severity was determined using the Ruuska and Vesikari scoring system [8]. The assessment was based on the following parameters: duration and maximum number of episodes of diarrhoea as well as vomiting, fever, and dehydration. Scores 0 – 5 was considered as mild, 6 – 10 as moderate, 11 – 15 as severe and ≥16 as very severe. To assess the impact of RV1 by age, infants were categorised into four age groups (<6 months, 6 – 11 months, 12 – 23 months and 24 – 59 months).

### 2.4 Rotavirus detection and genotyping

A 10-20% stool suspension in diluent buffer was prepared for each specimen and used to screen for the presence of group A rotavirus using a commercially available Enzyme immunoassay (Rotaclone®, Meridian Bioscience, Ohio, USA). Rotavirus dsRNA was extracted from all rotavirus-positive stool samples using the Viral RNA Mini-Kit (Qiagen, Hilden, Germany). The dsRNA was reverse transcribed to complementary DNA (cDNA) using random primers (Invitrogen, Paisley, UK) and reverse transcriptase enzyme (Superscript III MMLV-RT, Invitrogen, Paisley, UK) [9]. The cDNA was used to assign G genotype (G1, G2, G3, G4, G8, G9, G10, G11, G12) and P genotype (P[4], P[6], P[8], P[9], P[10], P[11], P[14]) using a multiplex heminested RT-PCR as described previously [5].

### 2.5 Statistical analysis

All statistical analyses were performed in the R environment for statistical computing, version 4.0.2 [10] and GraphPad Prism version 8. Vesikari score distributions were compared between pre-RV1 unvaccinated children, post RV1-unvaccinated children and post RV1-vaccinated children as well as between genotypes using non-parametric Kruskal-Wallis tests. Wilcoxon rank-sum tests were used to compare Vesikari scores between two groups. A linear regression model (*Vesikari score* = β_0_ + β_1_ * [post RV1-unvaccinated] + β_2_ * [post RV2-vaccinated] + ϵ) was used to estimate the change in Vesikari score that could be associated with receipt of RV1 and RV1 vaccine period. This model was adjusted for the Mid-Upper Arm Circumference (MUAC), age, weight, gender and receipt of the Bacillus Calmette–Guérin (BCG) and Pentavalent (diphtheria, pertussis, tetanus, and hepatitis B and *Haemophilus influenzae* type b) EPI vaccines. Both unadjusted and adjusted linear regression analysis were used to estimate Vesikari scores by considering G genotypes separately, P genotypes separately and combined G and P genotypes. For instance, the following unadjusted model was used to estimate effects of combined G & P genotypes: Vesikari = β_0_ + β_1_ * [post RV1-unvaccinated] + β_2_ * [post RV2-vaccinated] + β_3_ [genotype G2P[4]] +β_4_ [genotype G2P[6]] + β_5_ [genotype G12P[6]] +β_6_ [genotype G12P[8]] + β_7_ [post RV1-unvaccinated * genotype G2P[4]] + β_8_ [post RV1-unvaccinated * genotype G2P[6]] + β9 [post RV1-unvaccinated * genotype G12P[6]] + β_10_ [post RV1-unvaccinated * genotype G12P[8]] + β11 [post RV1-vaccinated * genotype G2P[4]] + β12 [post RV1-vaccinated * genotype G2P[6]] + β_13_ [post RV1-vaccinated * genotype G12P[6]] + β_14_ [post RV1-vaccinated * genotype G12P[8]] + ϵ. Residuals were computed for the purpose of model diagnostics: homoscedasticity and normality of residuals, linearity of the relationship between the independent and dependent variables.

### 2.6 Ethics

Ethical approval was obtained from the Research Ethics Committee of the University of Liverpool, Liverpool, UK (000490) and the National Health Sciences Research Committee, Lilongwe, Malawi (#867).

## 3. Results

### 2.1 Reduction in gastroenteritis severity following vaccination with RV1

The characteristics of the study participants included in this analysis are summarised in Table 1. In total, 3,159 children were recruited, of which 401 (12.7%) were recruited before RV1 introduction, whereas 2,758 (87.3%) were recruited after RV1 introduction. A total of 80.6% (2,224/2,758) of children recruited in the post-RV1 period were vaccinated. Thus, across the entire study period, 70.4% (2,224/3,159) of the children were vaccinated with RV1.

**Table 1.**
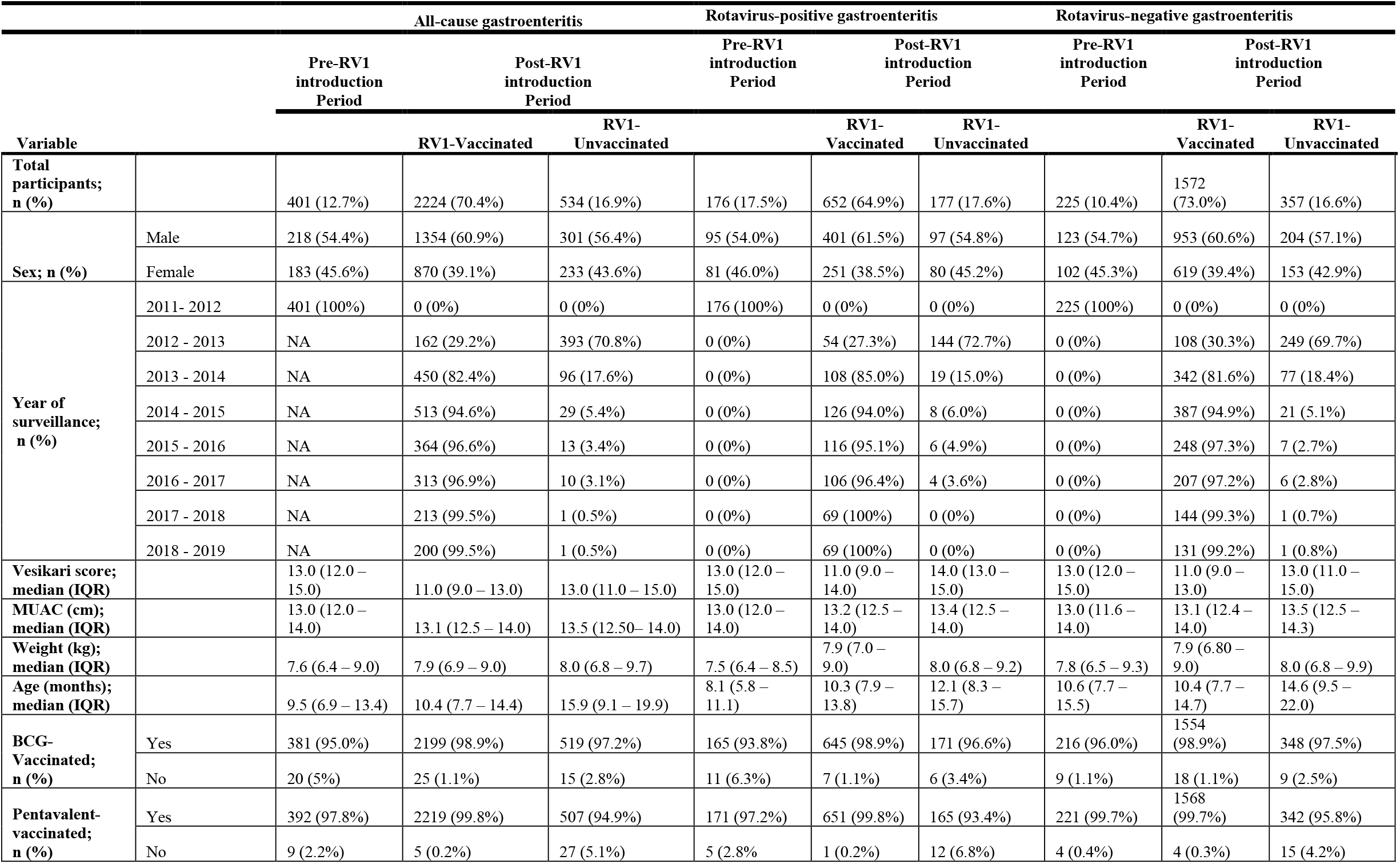
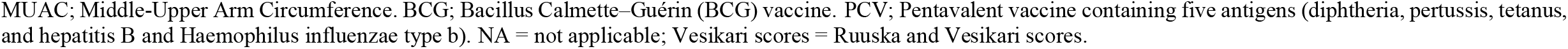
Characteristics of children who presented with gastroenteritis at the Queen Elizabeth Central Hospital in Blantyre, Malawi from December 2011 to October 2019.

RV1-vaccinated children presented with less severe gastroenteritis compared with RV1-unvaccinated children during the post-vaccine period (unadjusted Kruskal-Wallis test, *p* <0.001). There was no difference in the severity of gastroenteritis between RV1-unvaccinated children before and after RV1 introduction (unadjusted Wilcoxon rank-sum test, *p* = 0.260) (Figure 1a). When RV1-vaccinated children were stratified into rotavirus-positive and rotavirus-negative cases, a decrease in severity score was observed in RV1-vaccinated children for both groups (Figure 1b and 1c). Reductions in gastroenteritis severity three years or later following RV1 introduction were observed in all age groups (Figure 2 and Table S1).

**Figure 1.**
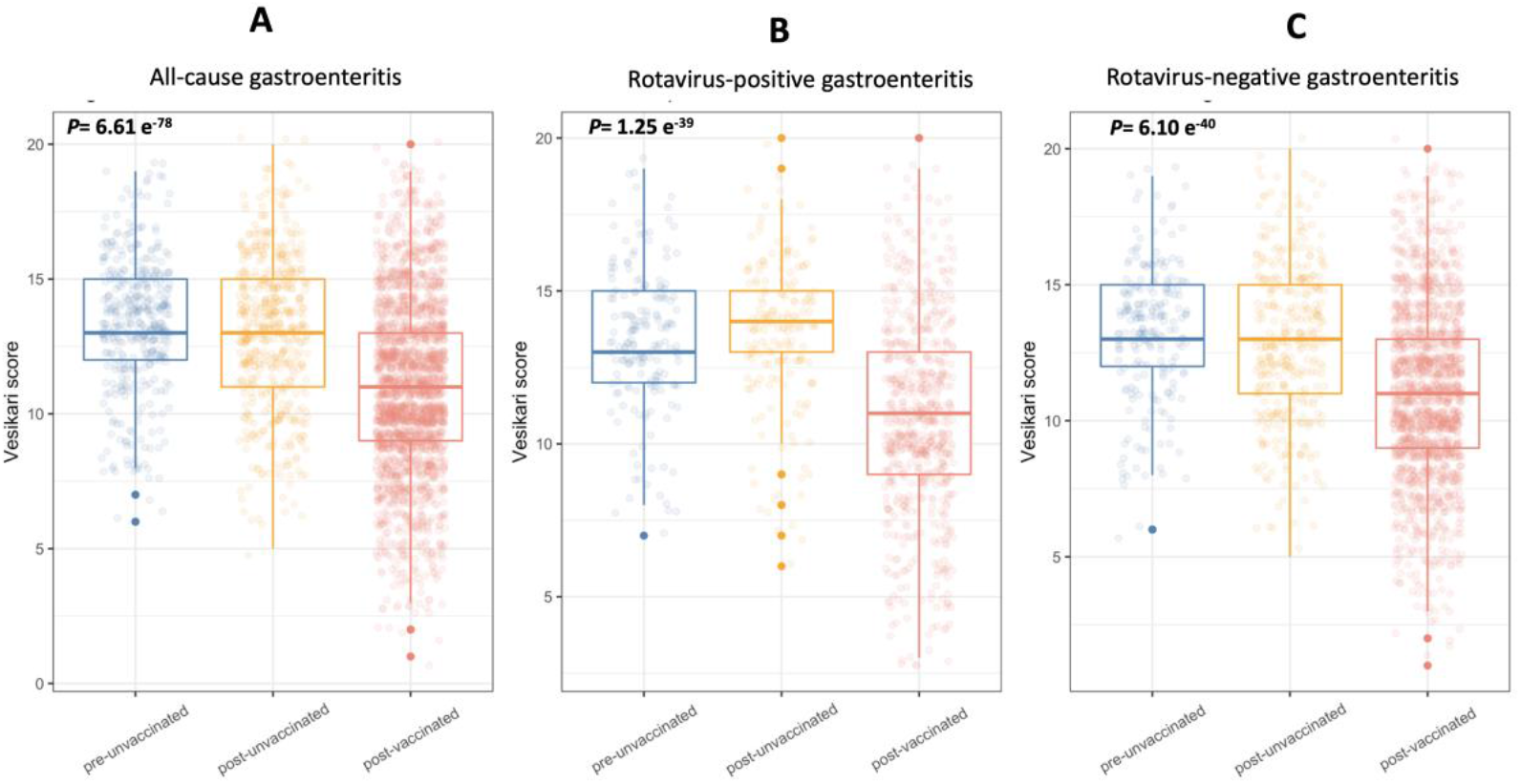
Ruuska and Vesikari severity scores among children hospitalized with gastroenteritis at QECH in Blantyre, Malawi before (December 2011 to October 2012) and after (November 2012 to October 2019) introduction of RV1. (A) Severity scores in all-cause gastroenteritis cases. (B) Severity scores in rotavirus-positive cases. (C) Severity scores in rotavirus-negative cases. Testing the null hypothesis that the severity scores have the same distribution in all 3 groups was highly statistically significant (*p*= 6.61e-78, 1.25e-39, 6.10e-40 for (A), (B) and (C), respectively; Kruskal-Wallis test). The *p*-value for observing the data under the null hypothesis of no different distribution between groups was 0.260 using a Wilcoxon rank-sum test when only comparing the pre-RV1 with the post-RV1 unvaccinated group (for all gastroenteritis cases). Vesikari scores = Ruuska and Vesikari score.

**Figure 2.**
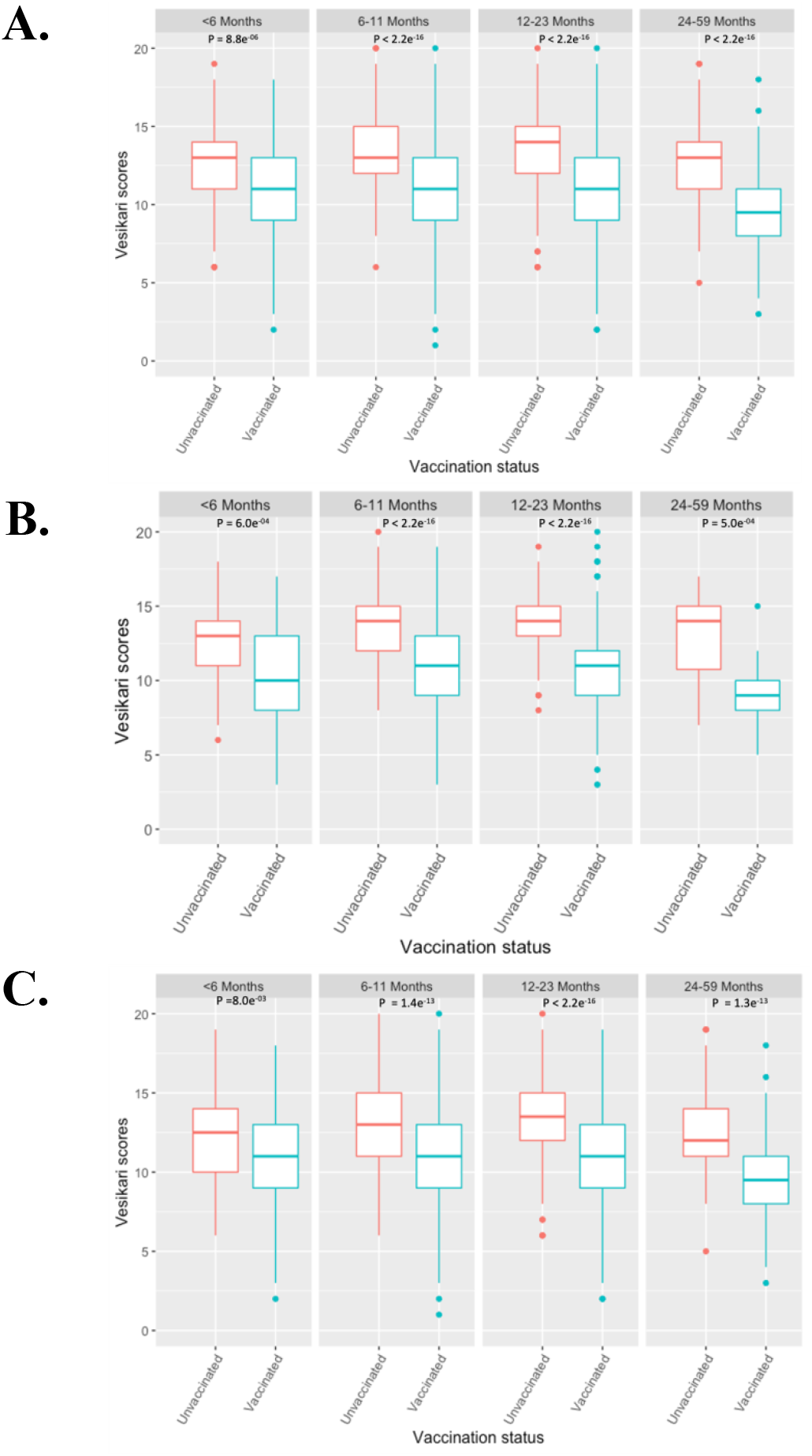
Comparison of Ruuska and Vesikari severity scores among children hospitalized with gastroenteritis at QECH in Blantyre, Malawi before (December 2011 to October 2012) and after (November 2012 to October 2019) introduction of RV1 stratified by age groups (Kruskal-Wallis test). (A) Severity scores in all-cause gastroenteritis cases (RV1-vaccinated, *n* = 248, 1128, 712 and 136; RV1-unvaccinated, *n*=121, 386, 311 and 117 in less than 6, 6 – 11, 12 – 23 and 24 – 59 months old children). (B) Severity scores in rotavirus-positive cases (RV1-vaccinated, *n* = 61, 353, 213, 25; RV1-unvaccinated, *n* = 69, 156, 112 and 16 in less than 6, 6 – 11, 12 – 23 and 24 – 59 months old children). (C) Severity scores in rotavirus-negative cases (RV1-vaccinated, *n* = 187, 773, 498 and 110; RV1-unvaccinated, *n* = 52, 225, 198 and 101 in less than 6, 6 – 11, 12 – 23 and 24 – 59 months old children).

Unadjusted regression analysis confirmed a linear relationship between the reduction in gastroenteritis severity and vaccination with RV1 when pre-RV1 unvaccinated hospitalised gastroenteritis cases were used as a reference group. There was an average estimated reduction of 2.35 (95% confidence interval (CI) 2.03, 2.67; *p*<0.001) in severity scores among RV1-vaccinated children and no reduction among RV1-unvaccinated children during the post-vaccine period (0.22; 95% CI -0.16, 0.61; *p*=0.260). Adjusting for the MUAC, age, weight, gender, and EPI vaccination status in the linear regression did not substantially change the estimated reduction in severity scores among RV1-vaccinated (2.21; 95% CI: 1.85, 2.56; *p*<0.001) and RV1-unvaccinated children (0.05; 95% CI -0.46, 0.36; *p*=0.820] (Table S2). Unlike the other covariates, there was some evidence of a linear association between age and gastroenteritis severity when children recruited before RV1 introduction and without a RV1 vaccination history were used as a reference group: every increment of 1 year in the age was associated with a decrease of 0.43 (95% CI 0.26, 0.60; *p*<0.001) in Ruuska and Vesikari score (Table S2). Except for < 6-month-old rotavirus-positive children where severity started to decline from the 2013 – 2014 calendar year, at least a year post-RV1 introduction, substantial decline in severity was observed in all age groups between 2014 and 2015, at least three years after RV1 introduction. Severity scores increased in all cases from 2017 to 2019 (Figure 2).

### 3.2 Relationship between rotavirus genotype, gastroenteritis severity and vaccination with RV1

The most frequently detected rotavirus genotypes were G1P[8], G2P[4], G2P[6], G12P[6] and G12P[8] [5], which comprised 66.57% (636/1,050) of all genotypes (Table S3). In RV1 unvaccinated children, regardless of the genotype, most gastroenteritis episodes were classified as severe (66.67%, 424/636) and severity scores did not differ by genotype (unadjusted Kruskal-Wallis test, *P*=0.544). In RV1-vaccinated children, a decrease in severity was observed for infections with all rotavirus genotypes compared to RV1-unvaccinated children, with most vaccinated children having moderate disease. This decrease in severity was less pronounced in cases infected with G12P[6] and G12P[8] genotypes; and RV1-vaccinated children infected with G12P[6] strains had a 2.58 unit (95% CI 0.60, 4.56; *p*=0.011) higher severity score when compared with the remaining strains after adjusting for age (Figure 3 and Table S4). Stratifying severity scores by G and P genotypes separately suggested that this effect might be associated with the G12 genotype (Figure S2). There were no differences in severity scores between pre-RV1 and post-RV1 unvaccinated groups in G and P genotypes, but a clear difference within the post-RV1 vaccinated group (Figure S2). The decrease in severity among post-RV1 vaccinated children was more pronounced for genotypes G1 and G2 compared to G12 (unadjusted Kruskal-Wallis test, *p* <0.001). Adjusting for age did not affect the regression outcomes (data not shown).

**Figure 3.**
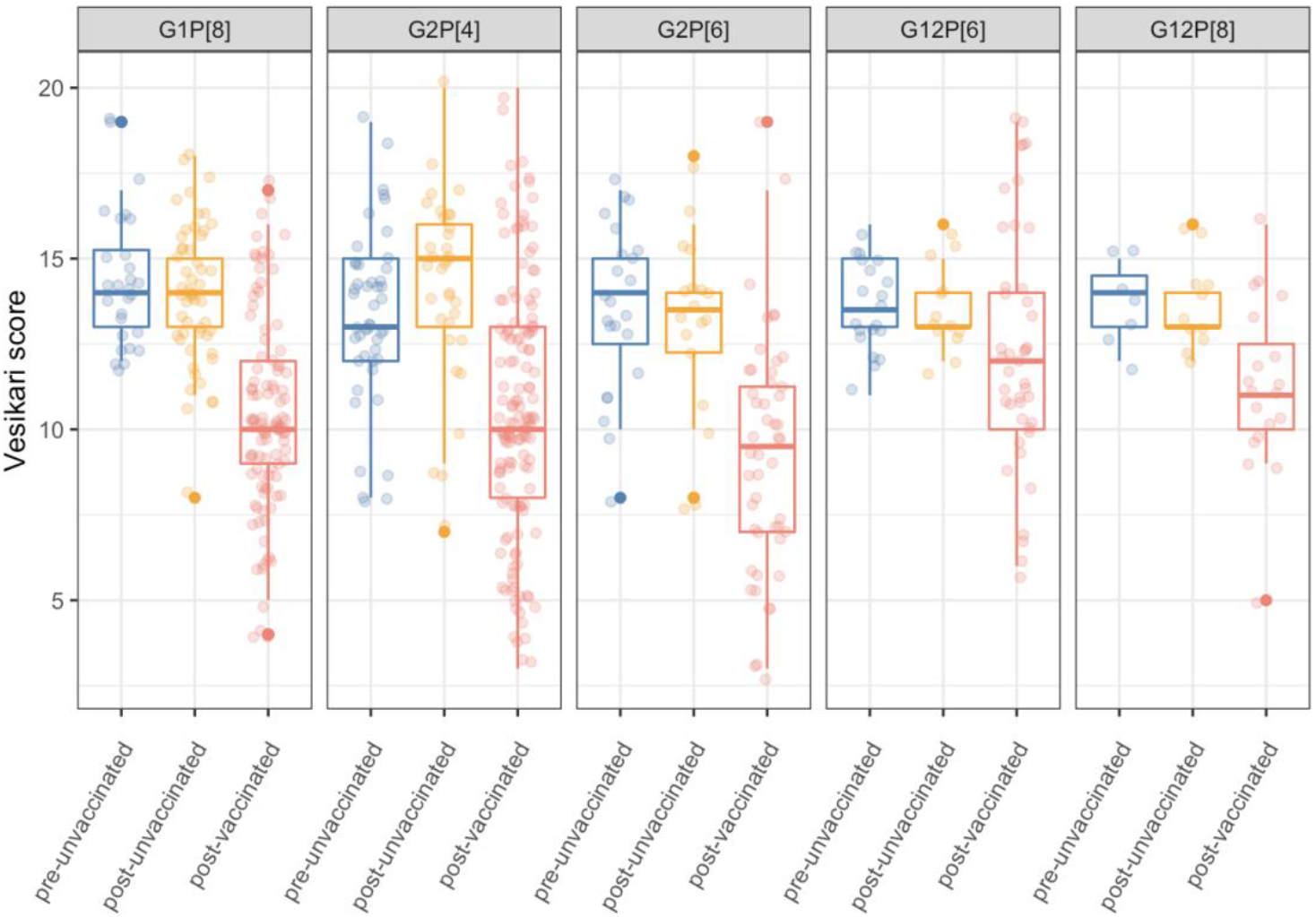
Comparison of severity scores in RV1-vaccinated and RV1-unvaccinated hospitalized children at Queen Elizabeth Central Hospital in Blantyre, Malawi presenting with gastroenteritis stratified by frequently detected combined G and P rotavirus genotypes. Vesikari scores = Ruuska and Vesikari scores.

## 5. Discussion

Introduction of RV1 into Malawi’s childhood immunisation schedule was associated with a significant reduction in the severity of all-cause gastroenteritis in children under the age of five years hospitalised at QECH in Blantyre. Irrespective of the rotavirus genotype, RV1-vaccinated children presented with less severe rotavirus disease compared to RV1-unvaccinated children during the pre- and post-RV1 introduction periods.

RV1 was developed to prevent children from developing severe gastroenteritis following infection with rotaviruses post-vaccination and not necessarily to prevent them from getting infected with rotaviruses [11, 12]. Thus, our study demonstrates expected, but to date unmeasured, direct impact of RV1 in reducing the severity of disease in children with acute rotavirus gastroenteritis in a low-income setting. The reduction in the severity of gastroenteritis in older children observed in the current study in both RV1-vaccinated and RV1-unvaccinated children could be explained by the potential acquired rotavirus-specific immunity either through rotavirus vaccination [6, 7, 13] or constant exposure to natural rotavirus infection due to the prevailing high force of rotavirus infection in Malawi [14], as the children grow older.

We also identified unexpected off-target vaccine benefit as RV1 recipients presented with less severe gastroenteritis regardless of the presence or absence of rotavirus. This is consistent with previous findings in Malawi and other settings where introduction of RV1 was associated with a reduction in all-cause diarrhoea mortality [7, 15, 16]. In contrast, a study that assessed the performance of rotavirus and oral polio vaccines in Bangladeshi infants did not find any evidence of nonspecific effect of RV1 on diarrheal burden or aetiology for a short period of 18 – 39 weeks post vaccination [17]. In a separate study in the same Bangladeshi population, delayed dosing of RV1 was associated with reduction in the duration of all-cause diarrhoea; however, rotavirus gastroenteritis was the only pathogen for which disease presentation was significantly reduced after a one-year follow-up [18].

Nonspecific vaccine effects were previously observed in infants vaccinated with BCG [19, 20] followed by herpes simplex [21, 22], measles [23] and other live attenuated vaccines such as the trivalent oral polio vaccine [17, 24]. T cell mediated cross-reactivity and trained innate immunity are among the mechanisms that may explain the off-target vaccine benefit [25, 26]. Vaccination with replication-deficient mutant herpes simplex type 1 virus provides protection from bacterial pathogens through induction of more robust CD8+ T cell cytotoxicity, prolonged production of interferon-gamma (IFN-γ), and systemic activation of macrophages [21, 22], whereas BCG elevates innate immune markers, such as IFN-γ, tumor necrosis factor α, interleukin 1βeta (IL-1β) and IL-6 cytokines [27, 28]. Oral live attenuated rotavirus vaccines could potentially employ similar mechanisms as they have been shown to induce innate immune responses [29] and effectively replicate in the gut of vaccinated children, [30-32] which could trigger cross-reactive CD8+ T cell responses. In addition, it is possible that averting rotavirus diarrhoea or experiencing less severe gastroenteritis in RV1-vaccinated children, [13, 33], promotes a healthier gut compared to their RV1-unvaccinated counterparts. The impaired gut integrity and physiology that follows severe rotavirus infection [34], may render them more susceptible to infection and/or more severe disease caused by other enteropathogens. This may potentially contribute, in part, to the indirect effects of rotavirus vaccine previously reported as herd protection [15, 16, 35-37].

RV1-vaccination was associated with a reduction in rotavirus gastroenteritis severity regardless of the infecting rotavirus genotype. Although RV1 effectiveness was previously documented to be lower against genotype G2-associated gastroenteritis compared with disease associated with genotype G1 [5, 38, 39], the reduction in severity in cases associated with G1 and G2 genotypes was similar among RV1-vaccinated children in the current study. However, the decline in gastroenteritis severity was less pronounced among cases associated with the G12 genotype. While the reasons for this observation are unknown, a relatively lower RV1 effectiveness has been reported against some heterotypic rotavirus strains, including G12s in this population [38]. Intriguingly, form 2017, rotavirus and non-rotavirus gastroenteritis severity scores increased in all age groups, although they remained lower than among those in unvaccinated children (Figure S1). Such a trend may be explained by potential changes in the criteria for hospitalisation of children presenting with gastroenteritis at QECH or changes in health seeking behaviour and or access of the communities around Blantyre post 2017. Further investigations are warranted to understand the significance and causes of this trend.

We could not test the trend in reduction of severity between vaccinated and unvaccinated infants because the number of unvaccinated infants decreased during each consecutive year due to increase in RV1 coverage. In addition, our analysis was limited by the availability of gastroenteritis cases from only one surveillance year prior to RV1 introduction, A further limitation is that all children were hospitalised, and hence had at least moderately severe gastroenteritis. Future studies should also examine the benefits of rotavirus vaccination in children with less severe gastroenteritis treated as outpatients or at surrounding health care facilities. Finally, the observed non-specific reduction in all-cause gastroenteritis could not be attributed to individual diarrhoea pathogens as the stool specimens were not routinely screened for pathogens beyond rotavirus.

In conclusion, our study provides evidence of a reduction in gastroenteritis severity among hospitalised children who had been vaccinated with RV1 seven years following its introduction in Malawi’s immunization programme. Rotavirus vaccination reduced the severity of rotavirus gastroenteritis caused by both homotypic and heterotypic rotavirus strains. Furthermore, rotavirus vaccination decreased non-rotavirus gastroenteritis severity, suggesting important off-target vaccine effects. Overall, these data demonstrate previously unmeasured direct and indirect benefits of rotavirus vaccines in Malawian children, providing further support for their continued programmatic use.

## Data Availability

The data referred to in this manuscript is freely available upon request.

## Supplementary Materials

**Supplementary Data S1, Table S1**: Gastroenteritis severity based on Vesikari scores among children presenting with gastroenteritis at the Queen Elizabeth Central Hospital in Blantyre, Malawi between the pre-(December 2011 – October 2012) and post-vaccine (November 2012 – October 2019) introduction periods.

**Supplementary Data S2, Table S2**. Unadjusted and adjusted linear regression model for the estimated reduction in severity scores among hospitalised children at Queen Elizabeth Central Hospital in Blantyre, Malawi after RV1 introduction.

**Supplementary Data S3, Table S3**. Frequently detected rotavirus strains and their gastroenteritis severity scores in children presenting with gastroenteritis at Queen Elizabeth Central Hospital, Blantyre, Malawi during the pre- and post-vaccine period (December 2011 to October 2019).

**Supplementary Data S4, Table S4**. Linear regression model for estimated reduction in severity scores and genotype (G and P), adjusted for age, MUAC and EPI vaccination status. Participants from the pre-RV1 period, with genotype G1P[8] and no vaccinations are taken as the reference group.

**Supplementary Data 5, Figure S1**. Trends in gastroenteritis severity among all children who presented with gastroenteritis at Queen Elizabeth Central Hospital in Blantyre, Malawi during the pre-(December 2011 to October 2012) and post-RV1 (November 2012 to October 2019) periods amongst different age groups. (A) All-cause gastroenteritis. (B) Rotavirus-positive gastroenteritis. (C) Rotavirus-negative gastroenteritis.

**Supplementary Data 6, Figure S2**. Severity scores in hospitalised children with rotavirus confirmed gastroenteritis at Queen Elizabeth Central Hospital, Blantyre, Malawi (A) Vesikari scores in RV1-vaccinated and RV1-unvaccinated children when only rotavirus P genotypes were considered. (B) Vesikari scores in RV1-vaccinated and RV1-unvaccinated children when only rotavirus P genotypes were considered.

## Disclaimer

The funders had no role in the study design, data collection and interpretation, or the decision to submit the work for publication. The authors did not receive any financial support or other form of reward related to the development of the manuscript. Therefore, findings and conclusions in this report are those of the authors and do not necessarily represent the formal position of the funders. D.H, M.IG., NAC., and K.C.J are affiliated to the National Institute for Health Research (NIHR) Health Protection Research Unit in Gastrointestinal Infections at University of Liverpool, a partnership with Public Health England, in collaboration with University of Warwick. The views expressed are those of the author(s) and not necessarily those of the NIHR, the Department of Health and Social Care or Public Health England.

## Acknowledgements

We acknowledge the support of the laboratory staff at the Malawi-Liverpool-Wellcome Trust Clinical Research Programme, clinical research team and the study participants. We thank Dr Naor Bar-Zeev, Dr Aisleen Bennett and Dr Louisa Pollock for their role in the collection of clinical data and faecal samples.

## Author Contributions

M.I.G, N.A.C. and K.C.J conceived and designed the study. K.C.J. collected clinical data and stool samples. C.M., J.J.M., E.C., O.K., K.C.J. performed the laboratory work. J.J.M. M.Y.R.H., D.H., M.I.G., and K.C.J carried out the statistical analysis. J.J.M. and K.C.J. drafted the manuscript. J.J.M., M.Y.R.H., C.M., R.W., A.W.K., C.M.B., E.C, O. K., I.T.S., D.H., M.I.G., N.A.C. and K.C.J contributed to interpretation of the data and writing the manuscript. All authors have read and approved the final manuscript.

## References

1. Clark A, Black R, Tate J, et al. Estimating global, regional and national rotavirus deaths in children aged <5 years: Current approaches, new analyses and proposed improvements. PloS one 2017; 12(9): e0183392.

2. Iturriza Gomara M, Wong C, Blome S, Desselberger U, Gray J. Molecular characterization of VP6 genes of human rotavirus isolates: correlation of genogroups with subgroups and evidence of independent segregation. Journal of virology 2002; 76(13): 6596–601.

3. Estes MK, Greenberg HB. Rotaviruses. In: Knipe DM HP, Cohen JI, Griffin DE, Lamb RA, Martin MA, et al. Fields Virology. 6th ed ed. Philadelphia, PA: Wolters Kluwer/Lippincott, Williams and Wilkins, 2013:1347–401.

4. Doro R, Laszlo B, Martella V, et al. Review of global rotavirus strain prevalence data from six years post vaccine licensure surveillance: is there evidence of strain selection from vaccine pressure? Infection, genetics and evolution : journal of molecular epidemiology and evolutionary genetics in infectious diseases 2014; 28: 446–61.

5. Mhango C, Mandolo JJ, Chinyama E, et al. Rotavirus Genotypes in Hospitalized Children with Acute Gastroenteritis Before and After Rotavirus Vaccine Introduction in Blantyre, Malawi, 1997 - 2019. J Infect Dis 2020.

6. Bar-Zeev N, Kapanda L, Tate JE, et al. Effectiveness of a monovalent rotavirus vaccine in infants in Malawi after programmatic roll-out: an observational and case-control study. The Lancet infectious diseases 2015; 15(4): 422–8.

7. Bar-Zeev N, King C, Phiri T, et al. Impact of monovalent rotavirus vaccine on diarrhoea-associated post-neonatal infant mortality in rural communities in Malawi: a population-based birth cohort study. Lancet Glob Health 2018; 6(9): e1036–e44.

8. Ruuska T, Vesikari T. A prospective study of acute diarrhoea in Finnish children from birth to 2 1/2 years of age. Acta Paediatr Scand 1991; 80(5): 500–7.

9. Iturriza-Gomara M, Green J, Brown DW, Desselberger U, Gray JJ. Comparison of specific and random priming in the reverse transcriptase polymerase chain reaction for genotyping group A rotaviruses. Journal of virological methods 1999; 78(1-2): 93-103.

10. Team RC. R: A language and environment for statistical computing. R Foundation for Statistical Computing, Vienna, Austria. 2020.

11. Dennehy PH, Brady RC, Halperin SA, et al. Comparative evaluation of safety and immunogenicity of two dosages of an oral live attenuated human rotavirus vaccine. The Pediatric infectious disease journal 2005; 24(6): 481–8.

12. Dennehy PH. Rotavirus vaccines: an overview. Clin Microbiol Rev 2008; 21(1): 198–208.

13. Clark HF, Furukawa T, Bell LM, Offit PA, Perrella PA, Plotkin SA. Immune response of infants and children to low-passage bovine rotavirus (strain WC3). Am J Dis Child 1986; 140(4): 350–6.

14. Pitzer VE, Bennett A, Bar-Zeev N, et al. Evaluating strategies to improve rotavirus vaccine impact during the second year of life in Malawi. Sci Transl Med 2019; 11(505).

15. Paternina-Caicedo A, Parashar UD, Alvis-Guzman N, et al. Effect of rotavirus vaccine on childhood diarrhea mortality in five Latin American countries. Vaccine 2015; 33(32): 3923–8.

16. Richardson V, Hernandez-Pichardo J, Quintanar-Solares M, et al. Effect of rotavirus vaccination on death from childhood diarrhea in Mexico. N Engl J Med 2010; 362(4): 299–305.

17. Upfill-Brown A, Taniuchi M, Platts-Mills JA, et al. Nonspecific Effects of Oral Polio Vaccine on Diarrheal Burden and Etiology Among Bangladeshi Infants. Clinical infectious diseases : an official publication of the Infectious Diseases Society of America 2017; 65(3): 414–9.

18. Colgate ER, Haque R, Dickson DM, et al. Delayed Dosing of Oral Rotavirus Vaccine Demonstrates Decreased Risk of Rotavirus Gastroenteritis Associated With Serum Zinc: A Randomized Controlled Trial. Clinical infectious diseases : an official publication of the Infectious Diseases Society of America 2016; 63(5): 634–41.

19. Biering-Sorensen S, Aaby P, Napirna BM, et al. Small randomized trial among low-birth-weight children receiving bacillus Calmette-Guerin vaccination at first health center contact. The Pediatric infectious disease journal 2012; 31(3): 306–8.

20. Aaby P, Roth A, Ravn H, et al. Randomized trial of BCG vaccination at birth to low-birth-weight children: beneficial nonspecific effects in the neonatal period? J Infect Dis 2011; 204(2): 245–52.

21. Lauterbach H, Kerksiek KM, Busch DH, et al. Protection from bacterial infection by a single vaccination with replication-deficient mutant herpes simplex virus type 1. Journal of virology 2004; 78(8): 4020–8.

22. Barton ES, White DW, Cathelyn JS, et al. Herpesvirus latency confers symbiotic protection from bacterial infection. Nature 2007; 447(7142): 326–9.

23. Aaby P, Martins CL, Garly ML, et al. Non-specific effects of standard measles vaccine at 4.5 and 9 months of age on childhood mortality: randomised controlled trial. BMJ 2010; 341: c6495.

24. Lund N, Andersen A, Hansen AS, et al. The Effect of Oral Polio Vaccine at Birth on Infant Mortality: A Randomized Trial. Clinical infectious diseases : an official publication of the Infectious Diseases Society of America 2015; 61(10): 1504–11.

25. Netea MG, Joosten LA, Latz E, et al. Trained immunity: A program of innate immune memory in health and disease. Science 2016; 352(6284): aaf1098.

26. Blok BA, de Bree LCJ, Diavatopoulos DA, et al. Interacting, Nonspecific, Immunological Effects of Bacille Calmette-Guerin and Tetanus-diphtheria-pertussis Inactivated Polio Vaccinations: An Explorative, Randomized Trial. Clinical infectious diseases : an official publication of the Infectious Diseases Society of America 2020; 70(3): 455–63.

27. Jensen KJ, Larsen N, Biering-Sorensen S, et al. Heterologous immunological effects of early BCG vaccination in low-birth-weight infants in Guinea-Bissau: a randomized-controlled trial. J Infect Dis 2015; 211(6): 956–67.

28. Kleinnijenhuis J, Quintin J, Preijers F, et al. Bacille Calmette-Guerin induces NOD2- dependent nonspecific protection from reinfection via epigenetic reprogramming of monocytes. Proceedings of the National Academy of Sciences of the United States of America 2012; 109(43): 17537–42.

29. Gandhi GR, Santos VS, Denadai M, et al. Cytokines in the management of rotavirus infection: A systematic review of in vivo studies. Cytokine 2017; 96: 152–60.

30. Cowley D, Boniface K, Bogdanovic-Sakran N, Kirkwood CD, Bines JE. Rotavirus shedding following administration of RV3-BB human neonatal rotavirus vaccine. Hum Vaccin Immunother 2017; 13(8): 1908–15.

31. Li JS, Cao B, Gao HC, et al. Faecal shedding of rotavirus vaccine in Chinese children after vaccination with Lanzhou lamb rotavirus vaccine. Sci Rep 2018; 8(1): 1001.

32. Markkula J, Hemming-Harlo M, Vesikari T. Shedding of oral pentavalent bovine-human reassortant rotavirus vaccine indicates high uptake rate of vaccine and prominence of G-type G1. Vaccine 2020; 38(6): 1378–83.

33. Bernstein DI, Smith VE, Sander DS, Pax KA, Schiff GM, Ward RL. Evaluation of WC3 rotavirus vaccine and correlates of protection in healthy infants. J Infect Dis 1990; 162(5): 1055–62.

34. Estes MK, Cohen J. Rotavirus gene structure and function. Microbiol Rev 1989; 53(4): 410–49.

35. Bennett A, Pollock L, Jere KC, et al. Direct and possible indirect effects of vaccination on rotavirus hospitalisations among children in Malawi four years after programmatic introduction. Vaccine 2018.

36. Buttery JP, Lambert SB, Grimwood K, et al. Reduction in rotavirus-associated acute gastroenteritis following introduction of rotavirus vaccine into Australia’s National Childhood vaccine schedule. The Pediatric infectious disease journal 2011; 30(1 Suppl): S25-9.

37. Van Effelterre T, Soriano-Gabarro M, Debrus S, Claire Newbern E, Gray J. A mathematical model of the indirect effects of rotavirus vaccination. Epidemiol Infect 2010; 138(6): 884–97.

38. Bar-Zeev N, Jere KC, Bennett A, et al. Population Impact and Effectiveness of Monovalent Rotavirus Vaccination in Urban Malawian Children 3 Years After Vaccine Introduction: Ecological and Case-Control Analyses. Clinical infectious diseases : an official publication of the Infectious Diseases Society of America 2016; 62 Suppl 2: S213–9.

39. Jere KC, Bar-Zeev N, Chande A, et al. Vaccine Effectiveness against DS-1-Like Rotavirus Strains in Infants with Acute Gastroenteritis, Malawi, 2013-2015. Emerg Infect Dis 2019; 25(9): 1734–7.

