## Supplementary Data S1, Table S1 for "Reduction in severity of all-cause gastroenteritis requiring hospitalisation in children vaccinated against rotavirus in Malawi"

**Supplementary Data S1, Table S1.** Gastroenteritis severity based on Ruuska and Vesikari scores among children presenting with gastroenteritis at the Queen Elizabeth Central Hospital in Blantyre, Malawi between the pre- (December 2011 – October 2012) and post-vaccine (November 2012 – October 2019) introduction periods.

|  | **Vaccination period** | | | | | | **Comparison between vaccination periods** | | |
| --- | --- | --- | --- | --- | --- | --- | --- | --- | --- |
|  | **Pre-RV1^1^** | | **Post-RV1 Unvaccinated^2^** | | **Post-RV1 Vaccinated^3^** | | **Pre-RV1 vs Post-RV1 Unvaccinated** | **Pre-RV1 vs Post-RV1 Vaccinated** | **Post-RV1 Vaccinated vs Post-RV1 Unvaccinated** |
| **Age (Mons^4^)** | **Number of samples** | **Vesikari scores(x̄)** | **Number of samples** | **Vesikari scores(x̄)** | **Number of samples** | **Vesikari scores(x̄)** | ***p*-values** | | |
| **All gastroenteritis cases^5^** | | | | | | | | | |
| **<6** | 72 | 12.2 | 49 | 12.6 | 248 | 10.7 | 0.488 | < 0.001 | 0.058 |
| **6 – 11** | 203 | 13.5 | 183 | 13.2 | 1128 | 11.3 | 0.228 | < 0.001 | < 0.001 |
| **12 – 23** | 94 | 13.8 | 217 | 13.5 | 712 | 10.8 | 0.19 | < 0.001 | < 0.001 |
| **24 – 59** | 32 | 13.2 | 85 | 12.3 | 136 | 9.5 | 0.084 | < 0.001 | < 0.001 |
| **0 – 59** | 401 | 13.3 | 534 | 13 | 2224 | 11 | 0.194 | < 0.001 | < 0.001 |
| **Rotavirus positive gastroenteritis cases^6^** | | | | | | | | | |
| **<6** | 45 | 12.3 | 24 | 12.6 | 61 | 10.4 | 0.36 | 0.001 | 0.17 |
| **6 – 11** | 94 | 13.7 | 62 | 13.9 | 353 | 11.1 | 0.352 | < 0.001 | < 0.001 |
| **12 – 23** | 35 | 14.2 | 77 | 13.9 | 213 | 10.7 | 0.364 | < 0.001 | < 0.001 |
| **24 – 59** | 2 | 13.5 | 16 | 12.8 | 24 | 9.2 | 0.496 | 0.006 | < 0.001 |
| **0 – 59** | 176 | 13.4 | 177 | 13.6 | 652 | 10.9 | 0.421 | < 0.001 | < 0.001 |
| **Rotavirus negative gastroenteritis cases^7^** | | | | | | | | | |
| **<6** | 27 | 12 | 25 | 126 | 187 | 10.8 | 0.549 | 0.044 | 0.156 |
| **6 – 11** | 104 | 13.4 | 121 | 12.9 | 773 | 11.4 | 0.128 | < 0.001 | < 0.001 |
| **12 – 23** | 58 | 13.6 | 140 | 13.2 | 498 | 10.9 | 0.269 | < 0.001 | < 0.001 |
| **24 – 59** | 30 | 13.2 | 71 | 12.2 | 110 | 9.6 | 0.045 | < 0.001 | < 0.001 |
| **0 – 59** | 219 | 13.3 | 357 | 12.8 | 1568 | 11 | 0.046 | < 0.001 | < 0.001 |

^1^Before RV1 was introduced. ^2^ RV1-unvaccinated children during the RV1 period. ^3^RV1-vaccinated children during the RV1 period. ^4^Age in months. ^5^Compares severity of gastroenteritis between vaccination periods and vaccination status amongst different age groups for all gastroenteritis cases. ^6^Compares severity of gastroenteritis between vaccination periods and vaccination status amongst different age groups for rotavirus-related gastroenteritis cases.^7^Compares severity of gastroenteritis between vaccination periods and vaccination status amongst different age groups for non-rotavirus related gastroenteritis cases. Vesikari scores = Ruuska and Vesikari scores.
