## Supplementary Data 5, Figure S1 for "Reduction in severity of all-cause gastroenteritis requiring hospitalisation in children vaccinated against rotavirus in Malawi"

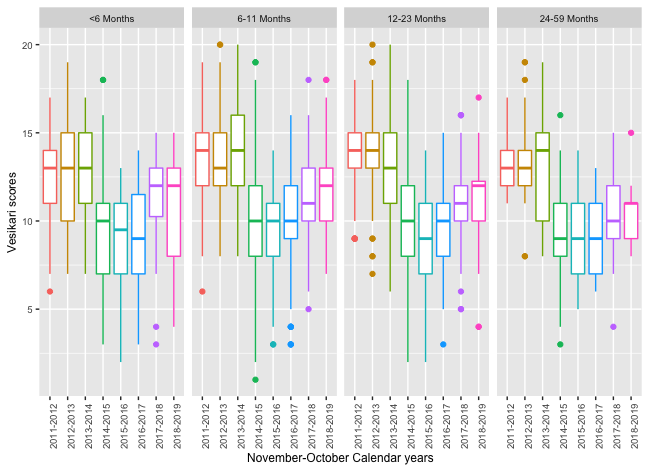

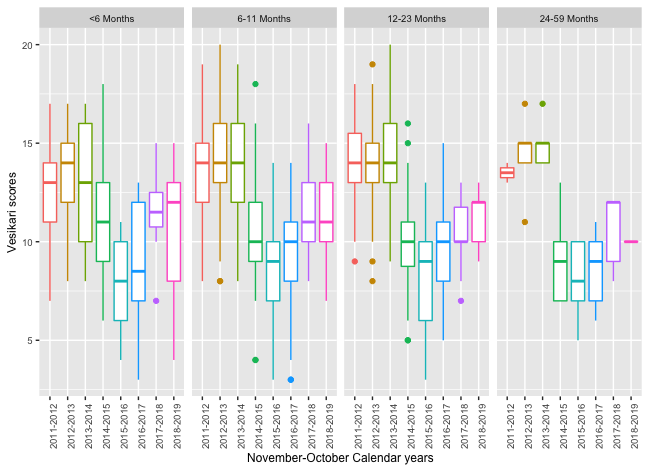

**C.**

**B.**

**A.**

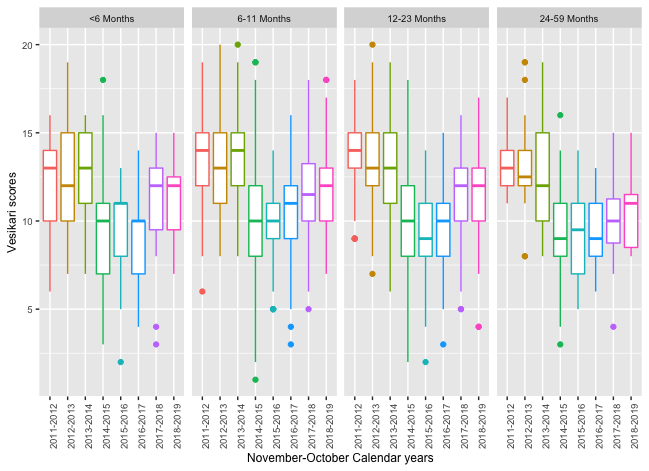

**Supplementary Data 5, Figure S1. Trends in gastroenteritis severity among all children who presented with gastroenteritis at Queen Elizabeth Central Hospital in Blantyre, Malawi during the pre- (December 2011 to October 2012) and post-RV1 (November 2012 to October 2019) periods amongst different age groups.** (A) All-cause gastroenteritis. (B) Rotavirus-positive gastroenteritis. (C) Rotavirus-negative gastroenteritis.
