## Supplementary Data 6, Figure S2 for "Reduction in severity of all-cause gastroenteritis requiring hospitalisation in children vaccinated against rotavirus in Malawi"

**B**

**A**

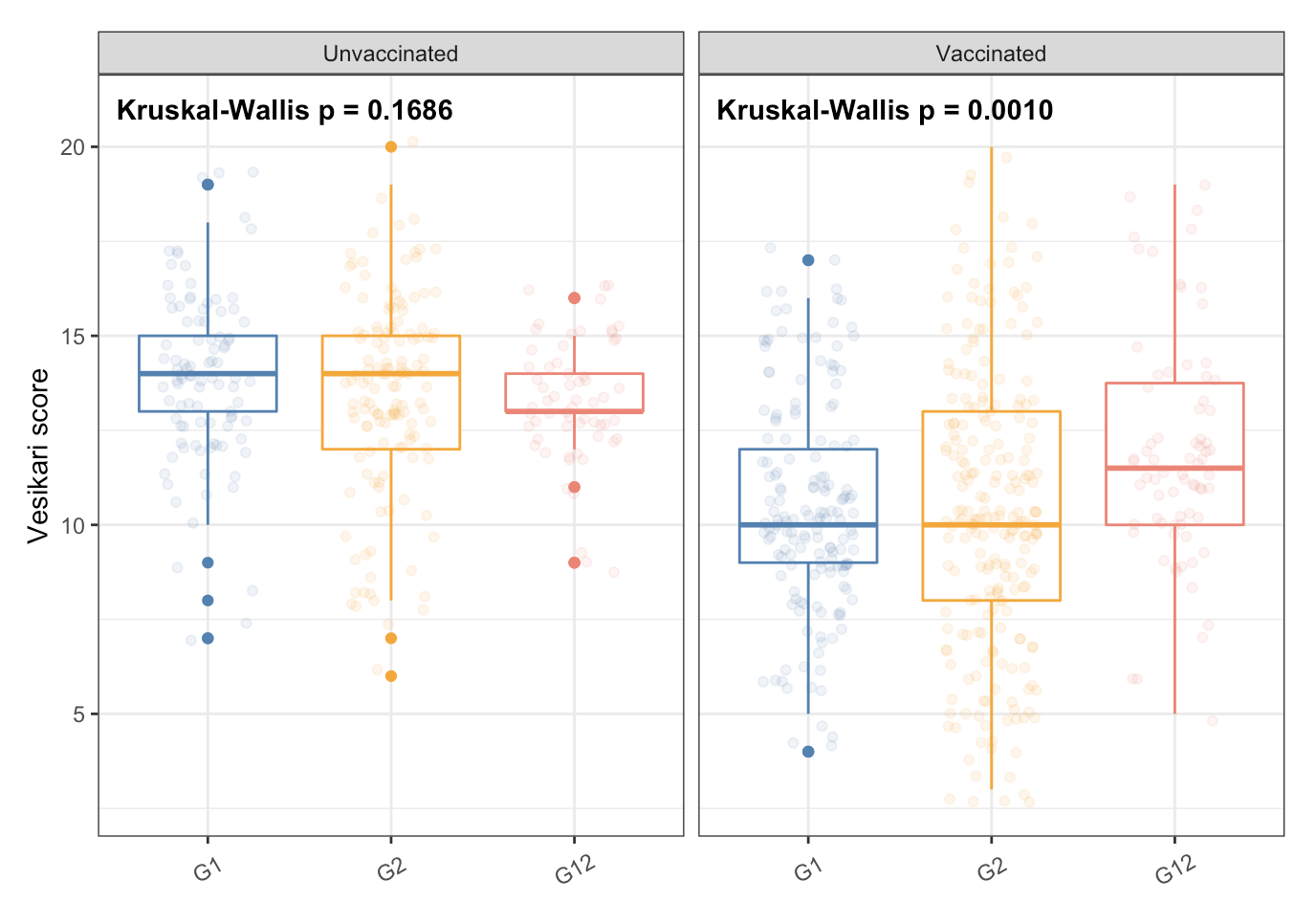

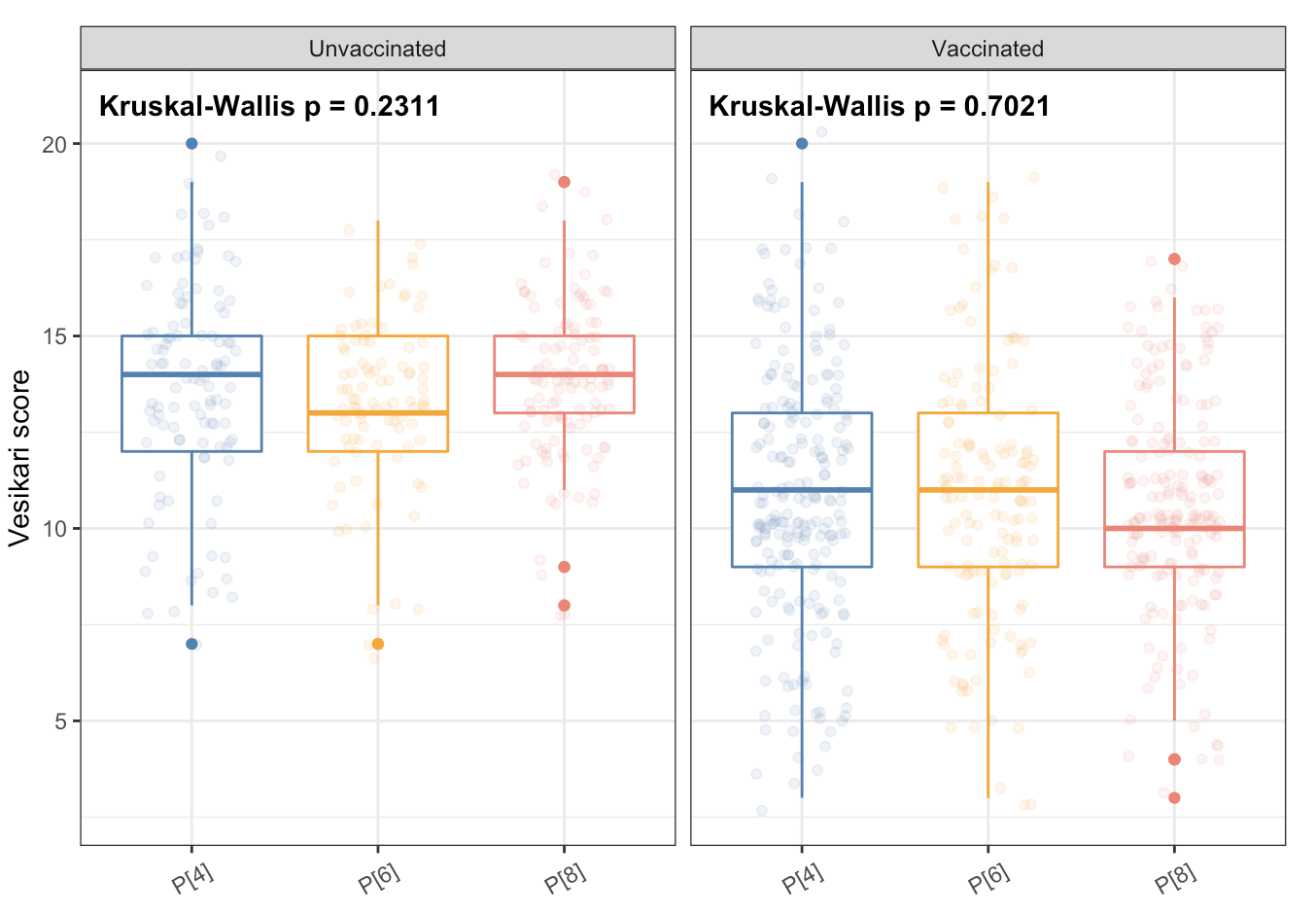

**Supplementary Data 6, Figure S2. Severity scores in hospitalised children with rotavirus confirmed gastroenteritis at Queen Elizabeth Central Hospital, Blantyre, Malawi** (A) Vesikari scores in RV1-vaccinated and RV1-unvaccinated children when only rotavirus P genotypes were considered. (B) Vesikari scores in RV1-vaccinated and RV1-unvaccinated children when only rotavirus P genotypes were considered. Vesikari scores = Ruuska and Vesikari scores.
