## Supplementary Data S2, Table S2 for "Reduction in severity of all-cause gastroenteritis requiring hospitalisation in children vaccinated against rotavirus in Malawi"

**Supplementary Data S2, Table S2.** Unadjusted and adjusted linear regression model for the estimated reduction in severity scores among hospitalised children at Queen Elizabeth Central Hospital in Blantyre, Malawi after RV1 introduction.

| **Variable** | **Coefficient estimate** | **95% confidence interval** | ***p*-value** |
| --- | --- | --- | --- |
| **Unadjusted ^a^** | | | |
| Intercept | 13.34 | 13.04 – 13.63 | <0.001 |
| Post-RV1 unvaccinated | -0.22 | -0.61 – 0.16 | 0.260 |
| Post-RV1 vaccinated | -2.351 | -2.67 – -2.03 | 0.001 |
| **Adjusted ^b^** | | | |
| Intercept | 13.08 | 11.92 – 14.24 | <0.001 |
| Post-RV1 unvaccinated | 0.05 | -0.36 – 0.46 | 0.821 |
| Post-RV1 vaccinated | -2.21 | -2.56 – 1.85 | <0.001 |
| Age | -0.04 | -0.05 – -0.02 | <0.001 |
| MUAC | 0.01 | -0.02 – 0.04 | 0.569 |
| BCG | -0.35 | -1.13 – 0.44 | 0.391 |
| Penta | 0.40 | -1.22 – 2.02 | 0.630 |
| Polio | 0.92 | -0.61 – 2.45 | 0.239 |
| PCV | -0.55 | -1.11 – 0.01 | 0.053 |

^a^ Unadjusted for other covariates. Pre-RV1 is taken as the reference group. Adjusted R^2^ = 0.10. ^b^ Adjusted for age, MUAC and vaccination status (BCG, penta, polio, PCV). Participants from the pre-RV1 period without any vaccinations were taken as the reference group. Adjusted R^2^ = 0.11. MUAC; Middle-Upper Arm Circumference. BCG; Bacillus Calmette–Guérin (BCG) vaccine. PCV; Pentavalent vaccine containing five antigens (diphtheria, pertussis, tetanus, and hepatitis B and Haemophilus influenzae type b).
