## Supplementary Data S3, Table S3 for "Reduction in severity of all-cause gastroenteritis requiring hospitalisation in children vaccinated against rotavirus in Malawi"

**Supplementary Data S3, Table S3**. Frequently detected rotavirus strains and their gastroenteritis severity scores in children presenting with gastroenteritis at Queen Elizabeth Central Hospital, Blantyre, Malawi during the pre- and post-vaccine period (December 2011 to October 2019).

| **Gastroenteritis severity associated with frequently detected rotavirus genotypes** | | | | | | | | | | |
| --- | --- | --- | --- | --- | --- | --- | --- | --- | --- | --- |
|  | **Mild** | | **Moderate** | | **Severe** | | **Very severe** | | **Total** | |
| **Genotypes** | **n** | **%** | **n** | **%** | **n** | **%** | **n** | **%** | **n** | **%** |
| **VP7 (G) Genotypes only^1^** | | | | | | | | | |  |
| **G1** | 4 | 1.6% | 93 | 36.0% | 129 | 50.0% | 32 | 12.4% | 258 | 100.0% |
| **G2** | 28 | 7.7% | 120 | 33.1% | 165 | 45.5% | 50 | 13.8% | 363 | 100.0% |
| **G12** | 1 | 0.7% | 28 | 20.7% | 91 | 67.4% | 15 | 11.1% | 135 | 100.0% |
| **Total** | **33** | **4.57%** | **241** | **31.88%** | **385** | **50.93%%** | **97** | **12.83%%** | **756** | **100.00%** |
| **VP4 (P) Genotypes only^2^** | | | | | | | | | |  |
| **P[4]** | 17 | 5.3% | 103 | 32.1% | 155 | 48.3% | 46 | 14.3% | 321 | 100.0% |
| **P[6]** | 7 | 3.0% | 67 | 28.8% | 134 | 57.5% | 25 | 10.7% | 233 | 100.0% |
| **P[8]** | 9 | 3.0% | 95 | 32.0% | 162 | 54.5% | 31 | 10.4% | 297 | 100.0% |
| **Total** | **33** | **3.88%** | **265** | **31.14%%** | **451** | **53.00%** | **102** | **11.99%** | **851** | **100.00%** |
| **Combined G and P Genotypes^3^** | | | | | | | | | |  |
| **G1P[8]** | 4 | 2.0% | 63 | 31.8% | 103 | 52.0% | 28 | 14.1% | 198 | 100.0% |
| **G2P[4]** | 17 | 7.5% | 69 | 30.5% | 104 | 46.0% | 36 | 15.9% | 226 | 100.0% |
| **G2P[6]** | 7 | 7.4% | 31 | 32.6% | 48 | 50.5% | 9 | 9.5% | 95 | 100.0% |
| **G12P[8]** | 1 | 2.6% | 7 | 18.4% | 27 | 71.1% | 3 | 7.9% | 38 | 100.0% |
| **G12P[6]** | 0 | 0.0% | 13 | 16.5% | 54 | 68.4% | 12 | 15.2% | 79 | 100.0% |
| **Total** | **29** | **4.46%** | **183** | **28.77%** | **336** | **52.83%** | **88** | **13.84%** | **636** | **100.00%** |

^1^ Numbers and proportions of four levels of gastroenteritis severity (Ruuska and Vesikari scores) for VP7 (G) genotypes only.

^2^ Numbers and proportions of four levels of gastroenteritis severity (Ruuska and Vesikari scores) for VP4 (P) genotypes only.

^3^ Numbers and proportions of four levels of gastroenteritis severity (Ruuska and Vesikari scores) for combined G and P genotypes.
