## Supplementary Data S4, Table S4 for "Reduction in severity of all-cause gastroenteritis requiring hospitalisation in children vaccinated against rotavirus in Malawi"

**Supplementary Data S4, Table S4.** Linear regression model for estimated reduction in severity scores and genotype (G and P), adjusted for age, MUAC and EPI vaccination status. Participants from the pre-RV1 period, with genotype G1P[8] and no vaccinations are taken as the reference group. Adjusted *R*^2^ = 0.24.

| **Variable** | **Coefficient Estimate** | **95% Confidence Interval** | ***p*-value** |
| --- | --- | --- | --- |
| Intercept | 14.53 | 11.77 – 17.29 | <0.001 |
| post-RV1 unvaccinated | -0.01 | -1.40 – 1.39 | 0.992 |
| post-RV1 vaccinated | -3.74 | -5.01 – -2.47 | <0.001 |
| Genotype G2P[4] | -0.99 | -2.37 – 0.39 | 0.159 |
| Genotype G2P[6] | -0.64 | -2.30 – 1.02 | 0.451 |
| Genotype G12P[6] ^1^ | -0.64 | -2.34 – 1.05 | 0.459 |
| Genotype G12P[8] | -0.37 | -2.82 – 2.07 | 0.765 |
| Age | -0.03 | -0.07 – 0.01 | 0.114 |
| MUAC | 0.02 | -0.02 – 0.06 | 0.361 |
| BCG | -0.37 | -1.97 – 1.24 | 0.654 |
| Penta | -0.66 | -3.87 – 2.56 | 0.690 |
| Polio | 1.50 | -0.95 – 3.95 | 0.232 |
| PCV | -0.82 | -1.98 – 0.34 | 0.169 |
| Post-RV1 unvaccinated: genotype G2P[4] | 1.20 | -0.68 – 3.07 | 0.212 |
| Post-RV1 vaccinated: genotype G2P[4] | 1.26 | -0.31 – 2.83 | 0.115 |
| Post-RV1 unvaccinated: genotype G2P[6] | -0.61 | -2.91 – 1.68 | 0.600 |
| Post-RV1 vaccinated: genotype G2P[6] | -0.53 | -2.47 – 1.41 | 0.591 |
| Post-RV1 unvaccinated: genotype G12P[6] | 0.06 | -2.40 – 2.53 | 0.960 |
| Post-RV1 vaccinated: genotype G12P[6] | 2.58 | 0.60 – 4.56 | 0.011 |
| Post-RV1 unvaccinated: genotype G12P[8] | -0.03 | -3.12 – 3.06 | 0.983 |
| Post-RV1 vaccinated: genotype G12P[8] | 1.29 | -1.54 – 4.12 | 0.373 |

^1^ RV1-vaccinated children had severity scores on average 3.74 (95% CI [2.47, 5.01]; p<0.0001) units lower than RV1-unvaccinated children, and while G12P[6] infections had on average severity scores 0.64 (95% CI [-1.05, 2.34]; p=0.4589) units higher compared to G1P[8] genotypes, this combined effect (average decrease of 3.74 – 0.64 = 3.10) was offset on average by an increase of 2.58 (95% CI [0.60, 4.56]; p=0.0111) units for children which were both RV1-vaccinated and G12P[6] infected. MUAC; Middle-Upper Arm Circumference. BCG; Bacillus Calmette–Guérin (BCG) vaccine. PCV; Pneumococcal conjugate vaccine. Penta: Pentavalent vaccine containing five antigens (diphtheria, pertussis, tetanus, and hepatitis B and Haemophilus influenzae type b).
